# Behind the Paywall: Unreported Financial Conflicts in Eminent Psychiatry Journals

**DOI:** 10.1101/2025.04.07.25325310

**Authors:** Francis J. Gesel, James H. Baraldi, Jessica L. Goldhirsh, Brian J. Piper

**Affiliations:** Department of Medical Education, Geisinger Commonwealth School of Medicine; Department of Anesthesiology and Critical Care, Perelman School of Medicine at the University of Pennsylvania; Center for Perioperative Outcomes Research and Transformation, Perelman School of Medicine at the University of Pennsylvania; Department of Psychiatry, Geisinger Medical Center; Center for Pharmacy Innovation and Outcomes, Geisinger

## Abstract

**Research Question:** To what extent do undisclosed financial conflicts of interest (COIs) exist among physician-authors in high-impact US-based psychiatry journals?

**Study Design:** Cross-sectional

**Objectives:** The study aimed to assess the prevalence and magnitude of undisclosed financial COIs within high-impact US-based psychiatry journals. The primary research question was determining the extent and distribution of financial COIs among physician-authors in these journals.

**Design:** This investigation analyzed financial COIs by comparing self-reported disclosures to journal(s) with payments mandatorily reported in the Open Payments database.

**Setting:** The study was conducted across two prominent US-based psychiatry journals: the *American Journal of Psychiatry* (*AJP*) and the *Journal of the American Medical Association Psychiatry* (*JAMA-PSY*), covering original research articles published from 1 January 2020 to 31 December 2022.

**Participants:** 2,872 research publications published from 1 January 2020 to 31 December 2022 were examined in *AJP* (n = 1,368) and *JAMA-PSY* (n = 1,504). Seventy-four original research articles authored by 27 physician-authors (*AJP* n=15, *JAMA-PSY* n=12) met the eligibility criteria.

**Interventions:** No interventions were conducted in this observational study.

**Primary and Secondary Outcome Measures:** Primary outcomes included total payments received by authors within the three years prior to publication and the proportion of undisclosed payments. Secondary outcomes assessed the payment types (research vs. general payments), demographic characteristics of authors, and study types associated with undisclosed COIs.

**Results:** US$4.54 million was paid to authors in the two journals, of which US$645,135 (14.2%) were undisclosed. *AJP* authors received US$205,943 (7.5% of total payments) in undisclosed payments, while *JAMA-PSY* authors received US$439,192 (24.8%). Research payments constituted 82.3% of all undisclosed payments. Total undisclosed payments among the top 10 highest-earning authors accounted for 84.8% (*AJP*) and 99.6% (*JAMA-PSY*) of all undisclosed payments to journals. Nearly all undisclosed payments, 96.2%, were made to authors conducting randomized controlled trials (RCTs).

**Conclusions:** Substantial undisclosed financial COIs were identified among the top 10 earners in high-impact psychiatry journals. These findings highlight potential risks to research transparency and integrity. Further research is needed to evaluate the effectiveness of disclosure policies and develop mechanisms to mitigate COIs in psychiatric research.

**Strengths and Limitations:**

- This is the first systematic study of financial conflicts of interest of physician-authors publishing original research in two of the highest impact factor psychiatry journals, the *American Journal of Psychiatry* and *JAMA Psychiatry*, suggesting relevance to influential research and clinical practice.
- The study employed data from OpenPaymentsData.cms.gov, an authoritative if incomplete source for study of financial conflicts of interest.
- The necessary stringency of subject eligibility criteria that make this study meaningful winnowed the domain of analysis from nearly 3,000 publications to fewer than 30 authors.

## Introduction

Conflicts of interest (COIs) in psychiatric research represent a persistent ethical challenge, heavily influenced by the intricate relationships between biomedical researchers and the pharmaceutical industry (1). These financial ties can unduly influence study outcomes, with industry-funded research frequently showing favorable results for the sponsor’s products compared to non-industry-funded studies (2). This bias is particularly concerning in psychiatry research contexts, where there is a lack of objective biomarkers for most mental health disorders (3–5). Additionally, the pursuit of academic promotion and professional recognition, often tied to publication quantity and impact, can contribute to COIs (6). When authors’ financial or professional interests compromise the ethical integrity of their publications, this conflict undermines the legitimacy of the research and its influence on clinical practice and professional advancement (7,8). Academic journals’ COI disclosure policies implemented in response to these concerns are often insufficient to address them fully. Instead, these measures merely may highlight the COIs without necessarily mitigating their impact (5). This interplay of incentives, financial and otherwise, with ethical responsibilities necessitates a robust and transparent approach to managing COIs to maintain public trust and ensure the reliability and validity of psychiatric research and practice (9).

In psychiatric research, COIs often manifest as the substantial financial relationships between academic researchers and pharmaceutical companies (10). It is well established that these financial ties can lead to biased research outcomes favoring the sponsors’ products (10,11, 11+). Additionally, the influence of pharmaceutical companies extends beyond research to clinical practice and medical education, potentially altering prescribing behaviors and the content of educational programs (12). Despite continued attention to financial COIs in psychiatric research, the prevalence and extent to which financial COIs exist within prominent US-based psychiatric research journals is incompletely understood (13,14)

For decades, the *American Journal of Psychiatry* (*AJP*) and the *Journal of the American Medical Association Psychiatry* (*JAMA-PSY*) have been among the most influential US-based research journals in the field of psychiatry, driving research directions, clinical practice, and healthcare policies (15,16). Given these journals’ authoritative status, financial COIs with possible relevance to articles published within them warrant further investigation (17). *JAMA-PSY* and *AJP* have policies requiring authors to disclose financial relationships from the past three years. *JAMA-PSY* focuses on relevant activities outside the submitted work, while *AJP* mandates disclosure regardless of relevance to the study’s subject (18,19). However, the effectiveness of these measures in fully addressing conflicts of interest remains unclear (20). Investigating the extent of financial COIs in these leading journals is crucial for ensuring the integrity of psychiatric research and maintaining public trust (2,7). Therefore, our study objectives were: 1) to delineate the prevalence and magnitude of financial COIs in the *AJP* and *JAMA-PSY*; 2) to investigate physician-author and study characteristics that are associated with financial COIs; and 3) to identify the companies providing payments that were undisclosed by authors and their relationships to the product under investigation.

## Methods

### Inclusion and Exclusion Criteria

Original research publications by US physician-authors (holding MD and/or DO degrees and whose affiliation was within the US) in the *AJP* (2022 Impact Factor = 19.2) and *JAMA-PSY* (Impact Factor = 25.9) from 1 January 2020 to 31 December 2022 were identified. *AJP* and *JAMA-PSY* request that authors report disclosures for the three years before submission. Articles categorized as non-original research by the journal(s) and those with the first and last author holding non-MD and/or DO degrees or whose affiliation was outside the US were excluded from the analysis. The first and last authors were identified in OpenPayments.CMS.Gov using their full names, specialties, and department affiliations.

### Data Collection

Total payments received by authors for the 36 months preceding their respective publication dates were identified using OpenPayments.CMS.GOV. Data included information on the author’s sex, specialty, and annual payment amounts of the main payment types (i.e., general and research payments) reported on OpenPayments. For each payment type, the company making the payment was identified and compared to the author’s (s’) self-reported disclosures to the journal(s). Undisclosed payments were identified as those made by companies not listed in the authors’ self-disclosures. Disclosed payments were identified by companies listed in the authors’ self-disclosures. Financial COIs were investigated by comparing authors’ self-reported disclosures to the journal(s) with their payments reported in OpenPayments.

### Statistics

GraphPad Prism (V.10.2.3) was used for statistical analysis and to create figures. The Mann-Whitney test assessed the difference between journal payment distributions. A 2×2 chi-square analysis was used to evaluate the association between disclosure status (i.e., undisclosed vs. disclosed) by the journals. A p < 0.05 was considered statistically significant. A flowchart was generated using Microsoft PowerPoint (V.2405).

## Results

### Article selection

Research publications (n = 2,872) that were published from 1 January 2020 to 31 December 2022 by physician-authors in the *AJP* (n = 1,368) and *JAMA-PSY* (n = 1,504) were examined. Among these publications, 352 articles in *JAMA-PSY* and 526 articles in *AJP* were categorized as original research by the respective journal. Of the 697 *JAMA-PSY* and 1,045 *AJP* authors of these publications, 139 in *JAMA-PSY* and 110 in *AJP* were eligible for OpenPayments profiles. Of these authors, 14 in *JAMA-PSY* and 17 in *AJP* had OpenPayments profiles. Two redundant author profiles existed in *JAMA-PSY* and *AJP*, leaving 12 and 15 unique OpenPayments profiles in *JAMA-PSY* and *AJP*, respectively (Figure 1).

**Figure 1.**
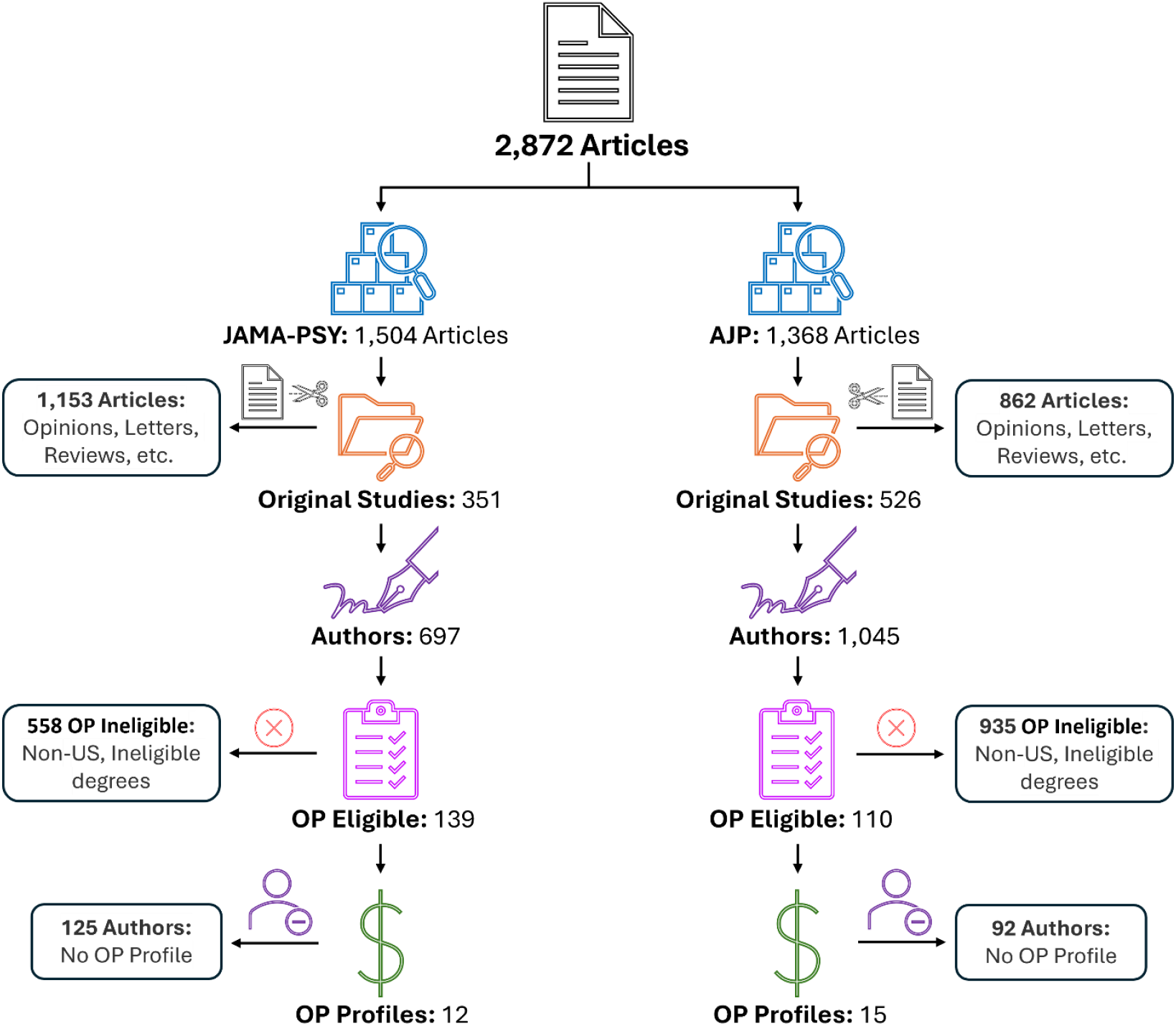
Flow chart illustrating the inclusion and exclusion criteria for physician-authors publishing in the *Journal of the American Medical Association Psychiatry* (*JAMA-PSY*) and the *American Journal of Psychiatry* (*AJP*) and as reported to CMS OpenPayments (OP).

### Total Payments

Combined payments to authors in AJP and JAMA-PSY were US$ 4,539,206.5 (19.7% GP US$ 895,000.0, 80.3% RP US$ 3,644,100.0). Authors received US$ 2,766,146.5 (9.0% GP US$ 248,600.0, 91.0% RP US$ 2,517,500.0) in AJP and US$ 1,773,060.0 (36.5% GP US$ 646,400.0, 63.5% RP US$ 1,126,600.0) in JAMA-PSY. Total mean payment (±SEM) amounts were US$ 18,722.0 (±US$ 9,176.0), GP US$ 17,759.0 (±US$ 5,962.0), RP US$ 359,646.0 (±US$ 223,226.0) in AJP and US$ 221,633.0 (±US$ 120,206.0), GP US$ 92,339.0 (±US$ 80,591.0), RP US$ 187,769.0 (±US$ 82,899.0) in JAMA-PSY.

The median payment for AJP authors was US$ 44,548.0 (GP US$ 8,140.0, RP US$ 144,204.0); at Q1 (25th percentile), the amount was US$ 4,571.0 (GP US$ 136.5, RP US$ 92,803.0), and at Q3 (75th percentile) the amount was US$ 213,813.0 (GP US$ 33,394.0, RP US$ 223,728.0). The median payment for JAMA-PSY authors was US$ 49,036.0 (GP US$ 2,395.0, RP US$ 108,353.0); at Q1 (25th percentile) the amount was US$ 1,138.0 (GP US$ 760.7, RP US$ 31,604.0) and at Q3 (75th percentile) the amount was US$ 448,194.0 (GP US$ 46,264.0, RP US$ 397,306.0).

### Undisclosed payments

Combined undisclosed payments of authors in AJP and JAMA-PSY accounted for 14.2% (US$ 645,135.7) of all payments. Of all undisclosed payments, 82.6% were research payments (US$ 532,841.1), and 12.6% were general payments (US$ 112,294.6) (p < 0.01). Undisclosed payments accounted for 7.5% (US$ 205,943.3, 16.9% GP US$ 34,808.8, 83.1% RP US$ 171,134.5) of total payments to AJP authors and 24.8% (US$ 439,192.4, 17.6% GP US$ 77,485.9, 82.4% RP US$ 361,706.6) of JAMA-PSY authors (Figure 2 and 3). The chi-squared analysis did not detect a significant association between undisclosed and disclosed payments between AJP and JAMA-PSY (p = 0.098).

**Figure 2.**
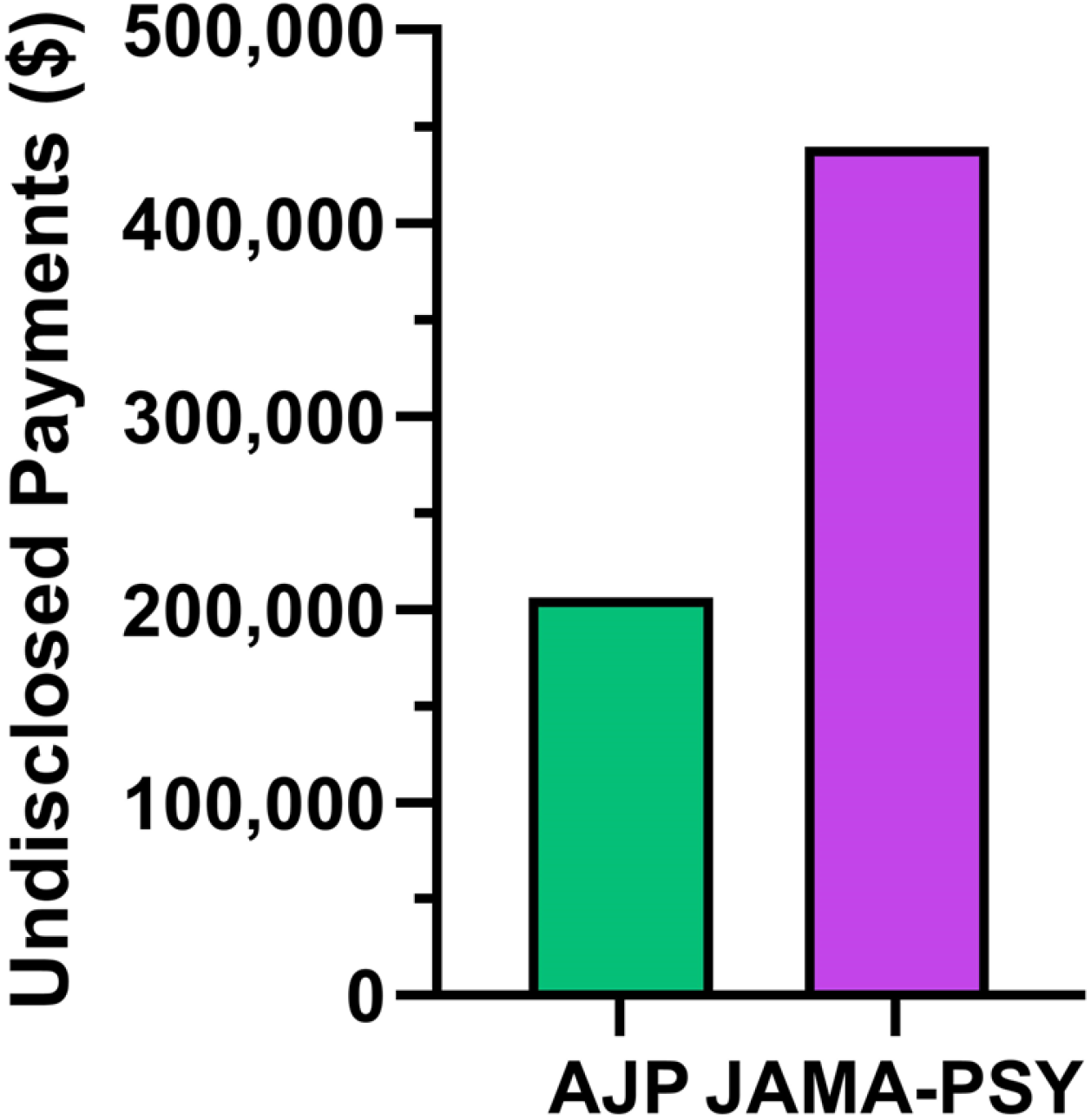
Total undisclosed payments among authors publishing in the *American Journal of Psychiatry* (*AJP*) and the *Journal of the American Medical Association Psychiatry* (*JAMA-PSY*).

**Figure 3.**
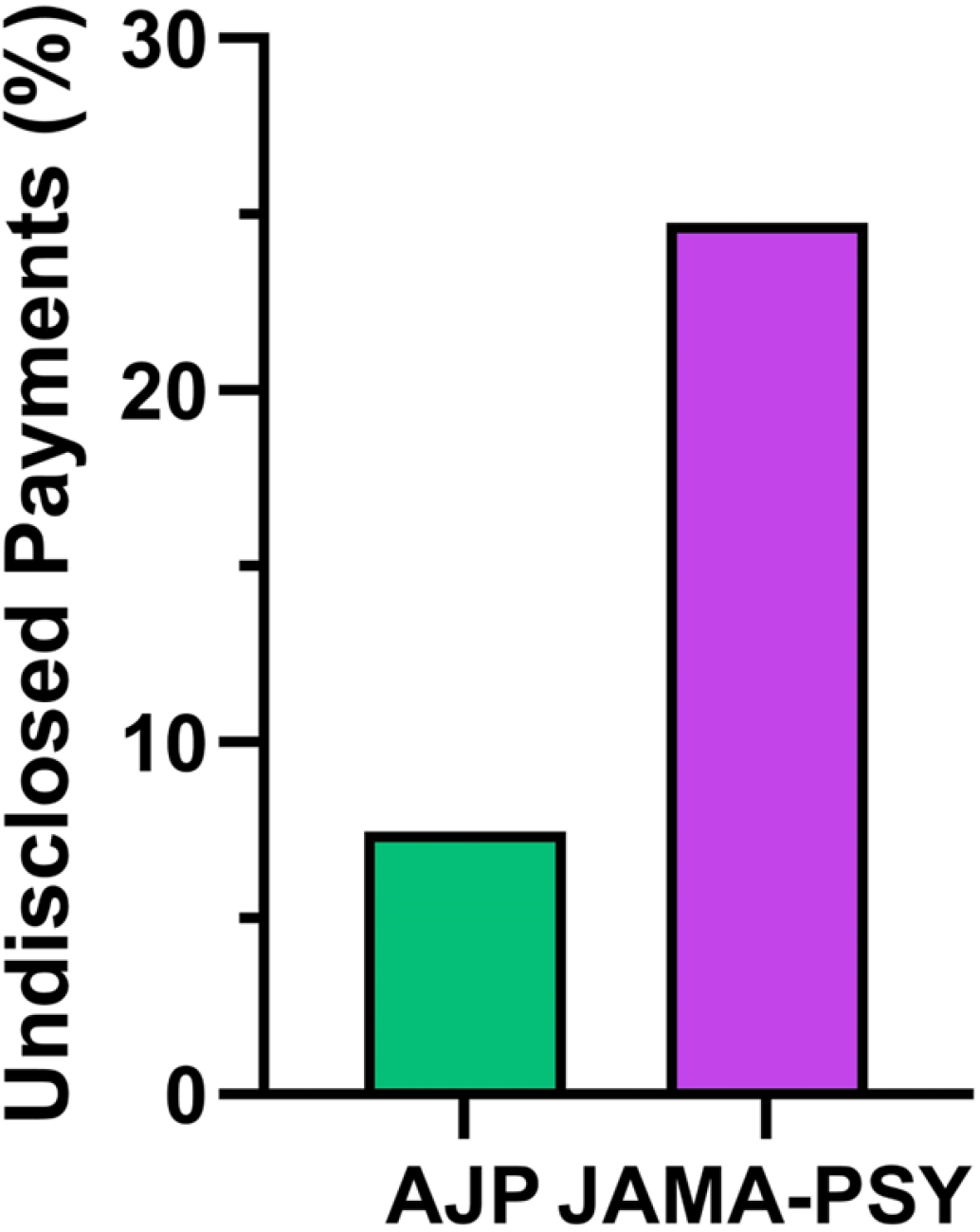
Percent of authors’ total compensation that was undisclosed by journal. *AJP*: American Journal of Psychiatry, *JAMA-PSY*: JAMA Psychiatry.

The median undisclosed payment amount for AJP authors was US$ 4,358.0 (GP US$ 2,008.0, RP US$ 38,627.0); at Q1 (25th percentile), the amount was US$ 144.8 (GP US$ 136.5, RP US$ 6,238.0) and at Q3 (75th percentile) the amount was US$ 21,548.0 (GP US$ 5,045.0, RP US$ 83,486.0). The median undisclosed payment amount for JAMA-PSY authors was US$ 26,917.0 (GP US$ 14,283.0, RP US$ 26,917.0); at Q1 (25th percentile), the amount was US$ 13,950.0 (GP US$ 760.7, RP US$ 13,569.0) and at Q3 (75th percentile) the amount was US$ 192,188.0 (GP US$ 62,442.0, RP US$ 153,825.0). Total mean (±SEM) undisclosed payment amounts were US$ 18,722.0 (±US$ 9,176.0), GP US$ 4,191.0 (±US$ 2,064.0), and RP US$ 42,784.0 (±US$ 20,682.0) in AJP and US$ 87,838.0 (±US$ 43,067.0), GP US$ 25,829.0 (±US$ 18,718.0), and RP US$ 72,341.0 (±US$ 35,673.0) in JAMA-PSY.

### Authors

Twenty-two (81.5%) of the authors were male (86.7% AJP n = 13, 75.0% JAMA-PSY n = 9). Twenty-four (88.9%) specialized in psychiatry, one (3.7%) in pathology, one (3.7%) in internal medicine, and one (3.7%) in obstetrics and gynecology. Eighteen (66.7%) were authors of RCTs, two (7.4%) of cohort studies, one (3.7%) of an economic evaluation study, two (7.4%) of cross-sectional studies, one (3.7%) of a meta-analysis, and three (11.1%) of diagnostic studies. Ten (37.0%) held the first authorship (AJP n = 6, JAMA-PSY n = 4), and 17 (63.0%) held the last authorship (AJP n = 9, JAMA-PSY n = 8) positions. Authors’ degrees included eighteen with an MD (66.7%), one MD/MSc (3.7%), one MD/DSc (3.7%), three MD/MPH (11.1%), and four MD/PhD (14.8%).

Of the 27 authors, 6 (22.2%) received no payments. Among the 21 (77.8%) authors receiving payments, 13 (61.9%) were in AJP, and 8 (38.0%) were in JAMA-PSY, and their payments ranged from a minimum of US$ 111.7 to a maximum of US$ 1,730,927.6. Sixteen (59.3%) of these authors (73.1% AJP n = 11, 26.7% JAMA-PSY n = 5) received undisclosed payments ranging from a minimum of US$ 15.7 to a maximum of US$ 206,045.6.

Eight of the top 10 highest-earning authors were male (*AJP* n = 5, *JAMA-PSY* n = 3). All 10 specialized in psychiatry and authored RCTs. Three held first authorship (*AJP* n = 2, *JAMA-PSY* n = 1), seven held last authorship (*AJP* n = 4, *JAMA-PSY* n = 3), and degrees included eight MDs (*AJP* n = 5, *JAMA-PSY* n = 3), one MD/PhD (*AJP* n = 1), and one MD/MSc (*JAMA-PSY* n = 1) (Figure 4).

**Figure 4.**
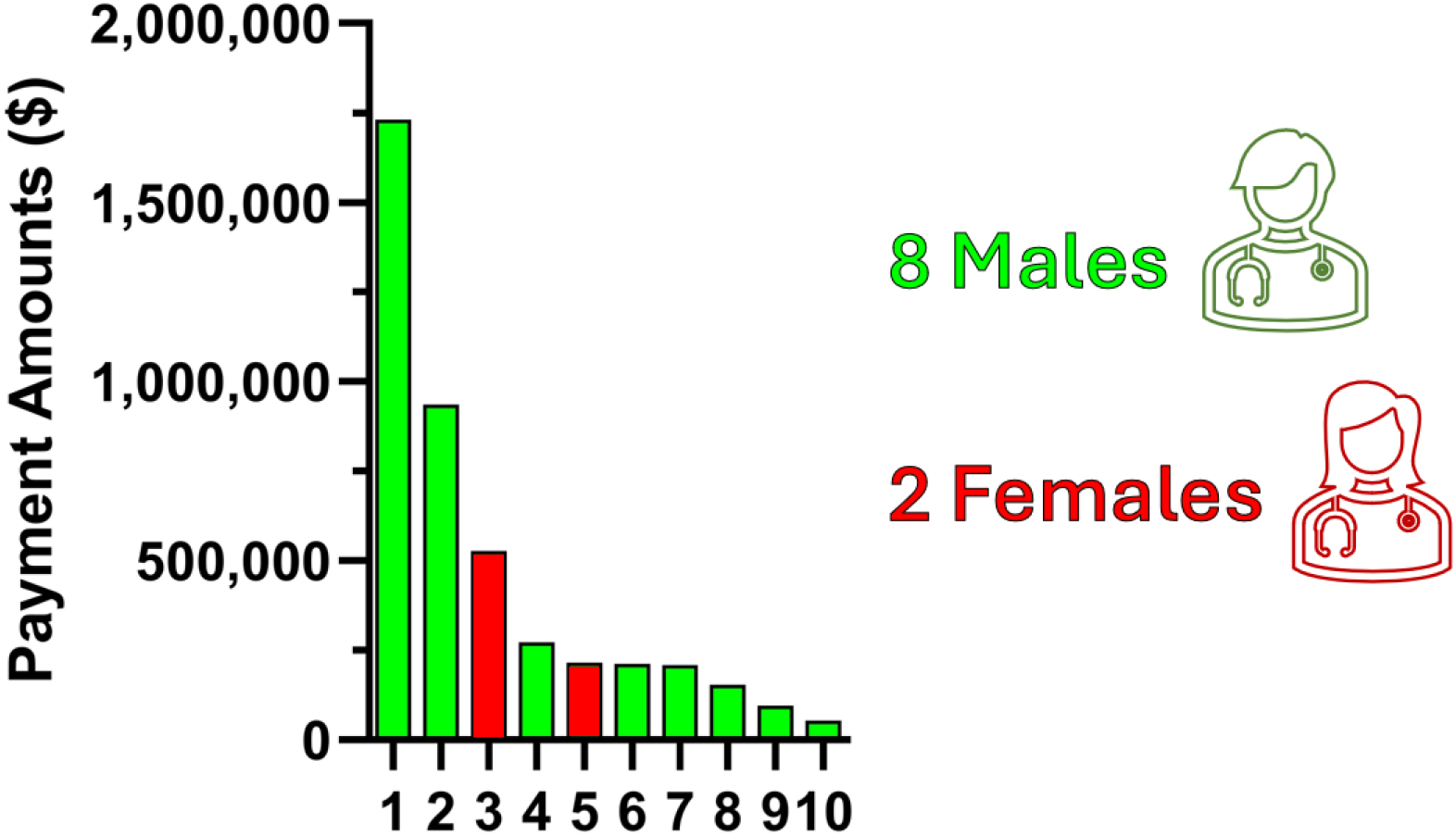
Study types and characteristics of the top 10 physician-authors publishing in the American Journal of Psychiatry and JAMA Psychiatry reporting the greatest total payments. All studies were randomized controlled trials except number eight which was a diagnostic report.

Their combined payments accounted for 76.5% (GP 27.6%, RP 88.5%), and their undisclosed payments were 94.9% (GP 9.5%, RP 99.0%) of all payments to the 27 authors. By journal, their combined payments represented 97.0% of all payments (GP 71.9%, RP 99.5%) to AJP and 44.6% to JAMA-PSY (GP 10.5%, RP 64.2%). Their combined undisclosed payments represented 84.8% of all payments (GP 22.5%, RP 97.4%) to AJP and 99.6% to JAMA-PSY (GP 99.0%, RP 99.8%).

#### Total Payments by Gender

Fifteen (68.2%) of the 22 males received payments totaling US$ 4,344,623.4 (95.7% of three-year total), US$ 888,893.6 (99.3% of general payments), and US$ 3,455,656.8 (94.8% of research payments). In contrast, all five (100.0%) of the females received payments amounting to US$ 194,583.1 (4.3% of the three-year total), US$ 6,101.8 (0.7% of general payments), and US$ 188,481.3 (5.2% of research payments).

### Undisclosed Payments by Gender

Thirteen (59.1%) of the 22 males received undisclosed payments totaling US$ 527,962.8 (81.8% of the three-year total), US$ 111,518.6 (99.3% of general payments), and US$ 416,444.2 (78.2% of research payments). Similarly, three (60.0%) of the five females received undisclosed payments amounting to US$ 117,172.9 (18.2% of the three-year total), US$ 776.1 (0.7% of general payments), and US$ 116,396.9 (21.8% of research payments).

### COI Disclosure Rates

Of the 27 authors who received payments, 12 made disclosures, the amount of which summed to at least half of the authors’ total payment amounts. Five had a three-year disclosure rate of 100%; two were published in *AJP*, and the other three were published in *JAMA-PSY*. The other eight authors who disclosed at least half of their payments had disclosure rates that ranged from 50.0% to 99.8%. Of the seven authors who disclosed less than half of their payment amounts ranged from 7.6% to 39.0%. One author disclosed 50.0% of her total payments. Six authors who received payments each disclosed 0%, or no amount, of their payments received. Of the authors who disclosed 0%, one published in *AJP*, and five published in *JAMA-PSY*.

### Undisclosed Payments by Study Topic & Company

The top 10 authors receiving the highest undisclosed payment amounts by study topic and company(−ies) are found in Table 1. Eight authors were male, and their combined undisclosed payments accounted for 81.7% of the total undisclosed payments received by all ten authors. The ten authors contributed to twelve RCTs, and eleven (91.7%) were pharmaceutical interventions. Seven (58.3%) studies focused on depression, including three using an antidepressant, one anticonvulsant, two antipsychotics, and one N-methyl-D-aspartate receptor (NMDAR) channel blocker for depression treatment. Other study topics included two (16.7%) on using a vasopressin 1a receptor antagonist for autism spectrum disorder, one (8.3%) on using an antidepressant medication for anxiety, one (8.3%) using cognitive behavioral therapy (CBT) for emotional and behavioral problems, and one (8.3%) genetic analysis on completed suicide. The mean (±SD) of the number of companies from which authors received payments but did not disclose was 2.4 (±2.2), with a range of one to eight.

**Table 1.** Characteristics of the top ten physician-authors publishing in the *American Journal of Psychiatry* (*AJP*) and the *Journal of the American Medical Association Psychiatry* (*JAMA-PSY*) with the greatest undisclosed payments as reported to CMS Open Payments, study topics, and companies providing payments to authors. All authors specialized in psychiatry. Percentage for companies indicates the amount of total undisclosed payments that were provided by the specific company General payments (GP), research payments (RP).

| Rank | Degree | Sex | Author ship | Journal | Total Undisclosed \$ | Undisclosed GP % | Undisclosed RP % | Companies |
| --- | --- | --- | --- | --- | --- | --- | --- | --- |
| 1 | MD | Male | First | *JAMA-PSY* | 206,045.6 | 66.0 | 37.9 | Genentech (US\$ 84,540.5, 41.0%), Neurocrine Biosciences (US\$ 83,856.7, 40.7%), Sunovion Pharmaceuticals (US\$ 27,687.7, 13.4%), Greenwich Biosciences (US\$ 4,264.1, 2.1%), Avanir Pharmaceuticals (US\$ 4,248.6, 2.1%), H. Lundbeck A/S (US\$ 3,196.7, 1.6%), Janssen Pharmaceuticals (US\$ 2,537.4, 1.2%), Boehringer Ingelheim Pharmaceuticals (US\$ 20.6, 0.0%) |
| 2 | MD | Male | Last | *JAMA-PSY* | 178,330.3 | 10.9 | 32.1 | Janssen Research & Development (US\$115,911.8, 65.0%), Lundbeck (US\$26,823.55, 15.05%), Pfizer (US\$35,594.92, 19.96%) |
| 3 | MD | Female | Last | *AJP* | 90,144.4 | 18.5 | 96.4 | Medtronic USA (US\$90,045.8, 99.8%), Boston Scientific Corporation (US\$98.6, 0.2%) |
| 4 | MD | Male | First | *AJP* | 65,504.4 | N/A | 29.3 | Boehringer Ingelheim Pharmaceuticals (US\$65,504.4, 100%) |
| 5 | MD, MSc | Female | Last | *JAMA-PSY* | 26,916.8 | 0.0 | 50.0 | Vanda Pharmaceuticals (US\$26,916.8, 100%) |
| 6 | MD | Male | Last | *JAMA-PSY* | 26,256.0 | 0.0 | 16.1 | Genentech (US\$26,256.0, 100%) |
| 7 | MD | Male | Last | *AJP* | 21,547.5 | 100.0 | 0.0 | Sage Therapeutics (US\$21,547.5, 100%) |
| 8 | MD, PhD | Male | Last | *AJP* | 11,750.0 | 0.0 | 8.3 | LivaNova USA (US\$11,750.0, 100%) |
| 9 | MD | Male | Last | *AJP* | 8,351.7 | 47.3 | 0.0 | SK Life Science (US\$4,393.93, 52.7%), Merck Sharp & Dohme (US\$1,949.3, 23.6%), ITI (US\$1,626.8, 19.5%), H. Lundbeck A/S (US\$381.6, 5.0%) |
| 10 | MD | Male | Last | *AJP* | 4,358.0 | 10.5 | 0.0 | Sage Therapeutics (US\$4,358.0, 100%) |

### Payments by Study Type

Three-year total payments to authors of RCTs accounted for 99.4% (US$ 4,513,573.7) of all payments, 97.2% (US$ 870,245.6) of general payments, and 100.0% (US$ 3,643,255.1) of research payments reported by all authors. Three-year total undisclosed payments to authors of RCTs accounted for 96.2% (US$ 620,472.9) of all undisclosed payments, 78.8% (US$ 88,514.8) of undisclosed general payments, and 99.8% (US$ 531,958.1) of undisclosed research payments reported by all authors.

In AJP, three-year total undisclosed payments to authors of RCTs accounted for 88.8% (US$ 182,924.2) of all undisclosed payments, 33.9% (US$ 11,789.7) of undisclosed general payments, and 100.0% (US$ 171,134.5) of undisclosed research payments reported by all authors. In JAMA-PSY, three-year total undisclosed payments to authors of RCTs accounted for 99.6% (US$ 437,548.7) of all undisclosed payments, 99.0% (US$ 76,725.1) of undisclosed general payments, and 99.8% (US$ 360,823.6) of undisclosed research payments reported by all authors.

Three-year total undisclosed payments to authors of cohort, economic evaluation, cross-sectional, meta-analysis, and diagnostic studies combined accounted for 3.8% (US$ 24,662.8) of all payments, 21.2% (US$ 23,779.8) of general payments, and 0.2% (US$ 883.0) of research payments reported by all authors.

### Undisclosed Payments by Company

The top ten companies providing the most payments that were undisclosed by authors and their corresponding percentage of the total undisclosed payments reported were Janssen Research & Development, LLC (US$ 115,888.0, 18.0%), Genentech, Inc. (US$ 106,522.4, 16.5%), Neurocrine Biosciences, Inc. (US$ 83,833.9, 13.0%), Medtronic USA, Inc. (US$ 67,370.4, 10.5%), Biohaven Pharmaceuticals, Inc. (US$ 58,511.3, 9.1%), Pfizer Inc. (US$ 35,595.1, 5.5%), Sunovion Pharmaceuticals Inc (US$ 27,683.2, 4.3%), Vanda Pharmaceuticals Inc. (US$ 26,916.8, 4.2%), Lundbeck LLC (US$ 26,847.2, 4.2%), and H. Lundbeck A/S (US$ 26,595.0, 4.1%).

### Undisclosed Payments by Year

Total undisclosed payments reported by year were: 2022 (US$ 123,539.8, 18.4%), 2021 (US$ 89,856.2, 13.4%), 2020 (US$ 78,340.5, 11.7%), 2019 (US$ 228,369.6, 34.1%), 2018 (US$ 70,629.4, 10.5%), and 2017 (US$ 79,099.5, 11.8%). Total percentages of undisclosed payments by journal and year ranged from 0.01% in 2017 to 18.4% in 2022 in AJP and 0.0% in 2022 to 34.1% in 2019 in JAMA-PSY.

## Discussion

The novel discoveries of this report were that 41.7% of authors in *JAMA-PSY* and 73.3% in *AJP* received undisclosed payments that were in contravention of journal policy. Notably, among these authors receiving undisclosed payments, 80.0% of those authors in *JAMA-PSY* and 36.3% of those in *AJP* received substantial undisclosed payments (> US$5,000) according to the National Institutes of Health threshold for a Significant Financial Interest) (21). These findings help elucidate the prevalence and magnitude of financial COIs within two of the most influential US-based journals in psychiatry.

Undisclosed financial COIs are particularly concerning as they may undermine the validity of psychiatric research (13). In our study, 14.2% of the total payments in *AJP* and *JAMA-PSY*, amounting to $645,135.7, were undisclosed. These undisclosed payments predominantly comprised research payments (82.6%), with a smaller proportion being general payments (12.6%). The high prevalence of undisclosed payments suggests that existing disclosure policies are insufficient to ensure full transparency (22).

The findings from this study underscore the critical intersection between financial COIs and gender representation in psychiatric research. Male authors, who dominate senior authorship in high-impact journals, disproportionately accounted for most undisclosed financial payments reported. This aligns with broader trends of male-dominated leadership roles in academic psychiatry (23). While female researchers have made significant strides in first authorship and contribute prominently to research on psychosocial epidemiology and biological studies, their underrepresentation in leadership raises questions about systemic barriers and their implications for financial transparency (24–26). This study contributes to the conversation by highlighting the concentration of undisclosed financial COIs among male authors conducting RCTs, underscoring the need for more robust disclosure policies and equitable representation in clinical trials. Addressing these issues is essential to fostering transparency and inclusivity in psychiatric research, ultimately promoting integrity and public trust.

An association between the type of study conducted and its authors receiving undisclosed financial COIs was detected. Authors involved in RCTs accounted for 99.44% of total payments and 96.2% of undisclosed payments, emphasizing the considerable funding associated with these studies. The concentration of payments among authors of results of RCTs suggests that pharmaceutical companies prioritize these studies, which aligns with prior literature indicating that industry inquiry prioritizes studies investigating products with market potential (27). Moreover, our findings suggest stereotypical study characteristics most likely to receive financial COIs in psychiatric research, namely RCT studies authored by male psychiatrists published in high-impact psychiatry journals. Given the previously reported positive association between pharmaceutical industry-funded research and favorable study outcomes in psychiatric clinical trials, it is prudent that studies displaying such stereotypical characteristics receive scrutiny to detect potential financial COIs since they may ultimately influence clinical guidelines and treatment protocols in favor of industry objectives rather than to optimize patient care (28,29).

Our analysis of undisclosed payments by study topic and company highlights potential issues regarding financial COIs in psychiatric research. The ten authors who received the highest undisclosed payments collectively accounted for 81.7% of the total undisclosed payment amount reported by all twenty-seven authors. These authors predominantly conducted RCTs, with eleven studies (91.7%) focusing on pharmaceutical interventions. Among the study topics, seven out of twelve studies (58.3%) centered on depression, examining medications such as antidepressants, anticonvulsants, antipsychotics, and N-methyl-D-aspartate receptor (NMDAR) channel blockers. Other studies included treatments for autism spectrum disorder, anxiety, emotional and behavioral problems treated with cognitive-behavioral therapy (CBT), and genetic analysis related to completed suicide.

The undisclosed payments were predominantly sourced from major pharmaceutical companies, including Janssen Research & Development, LLC, Genentech Inc., and Neurocrine Biosciences Inc. For instance, one author received $206,045.6 in undisclosed payments for a study on balovaptan for childhood autism spectrum disorder, with considerable contributions from Genentech and Neurocrine Biosciences. Another notable case involved an author receiving $178,330.3 for a study on transdiagnostic modular CBT for youth emotional and behavioral problems, primarily funded by Janssen Research & Development (65.0%), Lundbeck (15.1%), and Pfizer (12.0%). Additionally, studies on CBT combined with duloxetine and escitalopram for major depressive disorder received substantial undisclosed payments from Medtronic USA (99.8%) and Boehringer Ingelheim Pharmaceuticals (100%). These undisclosed payments raise concerns about the potential bias in research outcomes, particularly in RCTs where industry funding can influence study design and interpretation (30,31). On the other hand, of the top ten authors with undisclosed COI, no author neglected to disclose the company of the product they were studying which attests to at least partial compliance with the journals’ disclosure policies.

### Strengths and Limitations

One of the primary strengths of this study is its comprehensive approach to examining financial COIs in prominent psychiatric research journals. By analyzing data from a recent period (January 2020 to December 2022) and focusing on original research publications, the study provides a detailed and relevant snapshot of the current landscape of financial COIs. Covering this period ensures the findings are up-to-date and reflective of current practices. Additionally, using data from OpenPayments provides an alternative approach to current journal disclosure policies to investigate financial relationships, which helps reveal financial COIs that may remain undetected. The study’s focus on undisclosed payments sheds light on gaps in the current disclosure system, providing an additional perspective on the effectiveness of existing policies in psychiatric research. This insight is invaluable for policymakers, journal editors, and researchers aiming to improve transparency and integrity in psychiatric research. However, the study is not without limitations. The reliance on data from OpenPayments, which involves mandatory reporting by entities such as Pfizer and light verification by individual physicians, means the findings are subject to the accuracy and completeness of these records. Any errors or omissions in the database could influence the study’s results. Another limitation is the focus on only two journals, which, while highly influential, may not fully represent the broader field of psychiatric research. Further research is necessary to determine whether these findings generalize to less impactful psychiatry journals or journals in other countries. Furthermore, the study does not account for non-financial COIs, such as academic incentives or personal biases, which may also impact research outcomes.

## Conclusion

This study highlights the prevalence and magnitude of undisclosed financial COIs within two of the most influential US-based psychiatry journals. Substantial undisclosed financial COIs were identified among authors receiving payments, primarily concentrated in research payments and among a subset of highly compensated authors conducting RCTs. These findings underscore potential gaps in existing journal disclosure policies and their enforcement, which may adversely influence research transparency, integrity, and trust in psychiatric literature.

The reliance of leading journals on self-reported disclosures and the high prevalence of undisclosed financial COIs highlights the need for enhanced strategies to mitigate these conflicts since current disclosure policies have proved insufficient in both peer-reviewed literature and academic textbooks (13,32,33). To improve accuracy and accountability, future policies should consider incorporating independent cross-verification mechanisms, such as requiring links to OpenPayments profiles during manuscript submission. Addressing these issues is essential for promoting unbiased clinical decision-making, safeguarding psychiatric research credibility, and ensuring patient welfare prioritization over industry objectives.

## Author Contributions

- **Francis J. Gessel:** Primarily composed the manuscript, conducted the research, and analyzed data.
- **James H. Baraldi:** Assisted with data analysis and manuscript preparation.
- **Jessica Goldhirsh**: Assisted with manuscript preparation.
- **Brian J. Piper:** Principal Investigator (PI) oversaw the project and contributed to the manuscript preparation and data analysis.
- All authors have seen and approved the manuscript.

## Patient Consent

This study did not involve collecting personal medical information about any identifiable living individual, and such information was not part of the study design. Therefore, patient consent was not required.

## Data Availability

All data relevant to this study has been incorporated into the article. Further data may be made available upon reasonable request.

